# Inequalities in NHS Staff Support among those from Ethnic Minority and Migrant groups during the COVID-19 Pandemic

**DOI:** 10.1101/2025.03.19.25324242

**Authors:** Bethany Croak, Danielle Lamb, Sharon AM Stevelink, Rupa Bhundia, Juliana Onwumere, Brendan Dempsey, Pamela Almeida-Meza, Zoe Chui, Neil Greenberg, Rosalind Raine, Charlotte Woodhead, Stephani L Hatch, Rebecca Rhead

## Abstract

**Objectives:** During the COVID-19 pandemic, NHS staff support services aimed to support healthcare workers’ (HCWs) wellbeing, alongside informal support from colleagues and managers. However, bullying, harassment, and discrimination against HCWs from ethnic minority and migrant groups, along with low wellbeing support uptake, suggest disparities in workplace support. This study aimed to address the following research questions: 1) How does support programme use vary by ethnicity and migration status? 2) How does the perception of support from managers and colleagues vary by ethnicity and migration status?

**Methods:** This study analysed NHS CHECK survey data which examined the mental health and occupational outcomes (including support use) of HCWs during the COVID-19 pandemic across 18 Trusts in England. Data from 9,769 participants who completed the baseline survey (launched April 2020) and the six-month follow-up was analysed using descriptive statistics and binary logistic regression.

**Results:** HCWs from White Other (AOR 0.79; CI 0.64-0.99) and Asian ethnic groups (AOR 0.65; CI 0.57-0.74) were less likely to feel supported by their colleagues than White British HCWs. Similarly, those born outside of the UK and EU were less likely to feel supported by their colleagues than UK-born HCWs (AOR 0.70; CI 0.52-0.94). No variations in support programme use or support from managers were found across ethnicity or migration status.

**Conclusions:** The study suggests equitable formal support but identified critical disparities in perceived collegial support for HCWs during the COVID-19 pandemic. Improving workplace wellbeing should address the underlying social and structural factors that influence peer support and belonging.

- **What is already known on this topic** – Healthcare workers (HCWs) in the UK from ethnic minority and migrant groups are more likely than White British HCWs to experience abuse and discrimination from other staff. Therefore, they may be less likely to use workplace support, and feel less supported by their colleagues or manager. However, this has not been examined.
- **What this study adds** – The study indicates that formal support mechanisms for HCWs during the COVID-19 pandemic were generally equitable. However, it highlights significant disparities in perceived collegial support, with HCWs from some ethnic minority groups and HCWs born outside of the UK reporting lower levels of peer support compared to White British HCWs.
- **How this study might affect research, practice or policy** – These findings suggest that while structured support systems may be in place, the day-to-day experiences of workplace camaraderie and informal support vary considerably, underscoring the need for targeted interventions to foster a more inclusive and supportive work environment.

## BACKGROUND

During the early phases of the COVID-19 pandemic, 59% of healthcare workers (HCWs) in England met the threshold for a probable common mental disorder (CMD), such as anxiety or depression [1]. To support wellbeing, National Healthcare Service (NHS) staff were offered a wide variety of formal support interventions at a Trust and national level, including Wellbeing Hubs [2] (set up to provide HCWs with rapid access to free mental health support), dedicated relaxation spaces [3] and practical support such as free parking [4]. Informal support from managers and colleagues was also found to positively impact the mental health of HCWs during the pandemic [5].

A qualitative study on HCWs’ support needs during COVID-19 found peer support was the primary source of support [6]. However, some HCWs may have struggled to access it due to poor working relationships and prejudices among staff. During the COVID-19 pandemic, HCWs from Black and Mixed/Other ethnic groups were more likely than White British HCWs to experience bullying, harassment and abuse – hereafter referred to as abuse – from other staff [7]. This has been consistently reported by the NHS Workforce Race Equality Standard (WRES) since 2015 [8], which collects and monitors data across a series of key indicators of workforce equality. The latest WRES report indicated 17% of those from ethnic minority groups reported discrimination from a manager or colleague compared to 7% of staff from the White ethnic group. Staff from ethnic minority groups account for 24% of the NHS workforce [9], therefore, it is crucial they feel confident to access adequate support for workforce health, reduced sickness absences, and lower staff turnover.

Another group of HCWs who are at a heightened risk of discrimination and abuse from other staff is those HCWs born outside of the UK. HCWs from migrant groups were more likely than UK-born staff to experience discrimination from colleagues [10]. Similarly, international nurses from White ethic groups reported the highest level of abuse from patients and from other staff [8]. Migration status and ethnicity are closely linked, and HCWs born outside the United Kingdom (UK) may face disadvantages in workplace support.

Given that HCWs from an ethnic minority group, and those born outside of the UK are more likely to experience abuse, they may receive less support from colleagues and managers. One qualitative study found HCWs from an ethnic minority group felt unsupported by their managers during the COVID-19 pandemic, particularly when their managers were not from an ethnic minority group (i.e. White British) [11]. This may have multiple implications for mental health; workplace discrimination and harassment can harm mental well-being [7], those impacted may be less likely to benefit from the protective influence of colleague support and be less confident in reporting concerns of abuse to their manager. However, to our knowledge, there is no quantitative examination of support from colleagues or managers that allows us to understand the extent of this issue.

Formal support services and interventions were well used during the early phases of the COVID-19 pandemic, with 58% of HCWs in one study reporting engaging with at least one service (use of food donations not included in this estimate) [12]. However, UK HCWs reported mixed feelings about staff support, valuing practical support such as free parking but citing access barriers to psychological support such as stigma and lack of time [4]. There is some evidence of particularly low uptake of NHS Wellbeing Hubs by HCWs from Black and Asian ethnic minority groups, with some feeling these services were not equipped to address the impacts of racism and discrimination [13]. In contrast, HCWs born outside the UK found formal support, such as counselling and healthy eating provisions, helpful [14].

While early qualitative evidence suggests these groups may feel less supported and use staff support programmes less frequently, there is no quantitative research examining inequalities in workplace support during the COVID-19 pandemic. Although the COVID-19 pandemic has passed, it exacerbated existing and ongoing systemic issues, such as unprecedented waiting lists [15] and difficulties with retention and understaffing [16, 17]. Additionally, racial hostilities towards migrants are still prevalent, as demonstrated by the far-right riots in the UK in 2024 [18]; this led to the British Medical Association putting out guidance on how to support colleagues who had been affected [19]. Therefore, understanding if HCWs used support during the pandemic, a time when support needs were arguably very high, can inform how we identify inequalities and better support these staff going forward.

To address these issues and identify inequalities in access to workplace support in the NHS, this study analysed data from a large-scale survey of HCW mental health and wellbeing to answer the following research questions:

1. How does support programme vary by ethnicity and migration status?
2. How does the perception of support from managers and colleges vary by ethnicity and migration status?

## METHOD

NHS CHECK is a longitudinal cohort study comprising online surveys and semi-structured interviews [20], investigating HCW wellbeing during the COVID-19 pandemic and beyond (April 2020 – present). NHS employees (clinical and non-clinical) from 18 participating NHS trusts (Supplementary Material A) were eligible to take part (see Lamb et al. for details) [1].

The baseline survey opened in April 2020 and closed in January 2021 (n = 23,346; response rate 15%), with the second survey launched in October 2020 and closing in August 2021 (n = 10,671; response rate 45.5%). Data from participants who had completed baseline (T1) and the six-month follow-up (T2) were included in this study (n = 9,769). Of those who completed baseline, ∼45% completed the six-month follow-up. The surveys were split into a short and long version whereby after completing the first set of questions, participants were given the choice to complete a longer survey. All measures used in this analysis are from the short survey and response frequencies refer to the short surveys.

### Measures

Demographic and occupational information, including ethnicity, migration status, role, age and sex were collected at baseline. Ethnicity was captured using the 17 ethnicity categories from the 2011 UK Census. Due to low sample sizes, participant ethnicity was grouped into five categories: ‘White British’, ‘White Other’, ‘Black’, ‘Asian’ and ‘Mixed/Other’. For migration status, participants were asked to select one of three options, ‘born in UK’, ‘born in European Union (not UK)’ and ‘born elsewhere’. Participants were asked to specify their main role and this was combined into four job role categories: doctor, nurse, other clinical and other non-clinical.

#### Support Used

In the six-month follow-up survey (T2), participants were asked if they had used any national or local support services provided by NHS England or their local NHS Trust. Participants were asked to select yes/no to a list of support programmes. The list of support programmes in the survey was informed by discussions with participating NHS Trusts and the NHS CHECK Patient Public Involvement (PPI) group. The support included psychological support, initiatives by other organisations, such as retail discounts, and organisational support, such as team debriefs. A binary outcome variable was defined as *support used* if participants had used at least one support programme and *support not used* if they had not reported using any programme listed.

#### Feeling Supported

Participants were also asked: “Since the COVID-19 pandemic, how well do you feel supported by your manager?” and the same question about support from colleagues. Participants were asked to rate the extent to which they felt supported by their colleagues and by their manager on a 5-point scale anchored from: ‘not at all’, ‘a little bit’, ‘moderately’, ‘quite a bit’ to ‘extremely’.

Binary outcome variables were created. Participants who selected ‘not at all’ or ‘a little bit’ were categorised as *little / no support,* and those who selected ‘moderately’ or ‘quite a bit’ or ‘extremely’ were categorised as *supported*.

#### Mental Health

The 12-item general health questionnaire (GHQ-12), issued at T2, was used to estimate the prevalence of probable common mental disorders (CMDs), with a score of four or more indicating a probable CMD, using the 0-0-1-1 scoring method [22]. A threshold at this level has an estimated sensitivity of 69% and specificity of 88% [23]. The GHQ-12 has good internal consistency (Cronach α = 0.94) [24]. GHQ-12 total score (T2) was used to adjust for mental health difficulties for research aims one and two.

### Statistical Analysis

Secondary data analysis was performed on a sub-sample of the NHS CHECK cohort; participants who had completed the baseline survey and the six-month follow-up were included in this analysis (n = 9769). All data analyses were conducted using Stata (v17) [25].

The proportion of participants who reported they had used support programmes and who felt supported by their manager and colleagues was calculated. Two-level random effects logistic regression models accounted for the hierarchical data structure, with HCWs nested within NHS Trusts. Primary outcomes were support programme use (RQ1) and perceived support from managers and colleagues (RQ2). Ethnicity and migration status were the main exposures, with separate regression models for each of the outcomes (six unadjusted and six adjusted). Models were fitted using Stata’s ‘melogit’ command for multilevel logistic regression with random effects.

Previous research found that UK HCWs who were female, younger and had higher psychological distress were more likely to use staff support services during the pandemic [12]. Supplementary analysis (supplementary material B) also showed associations between support use, psychological distress (measured by GHQ-12) and manager/colleague support. Therefore, all regression models (reported as AOR) adjusted for age, sex and psychological distress. Additionally, as NHS CHECK cohort data found nurses had poorer mental health than other HCWs [1] and psychological distress is associated with support use [12], we adjusted for job role in all analyses.

Response rates for each Trust were calculated using aggregate-level population data, including information regarding the total number of Trust staff, age, sex, and ethnicity of staff. Response weights were generated using a raking algorithm based on age, sex, job role (a dichotomised variable with two categories: clinical/non-clinical) and ethnicity and were implemented using the ‘svyset’ command [26]. Reported frequencies are unweighted, and proportions are weighted [25].

## RESULTS

### Data Completeness

Missing data for exposure, outcome and predictor variables is detailed in Table 2, the CMD variable is reported in table 3. As the proportion of missing data on each individual predictor and outcome variables was no more than 5%, complete case analysis was deemed appropriate [27]. For all adjusted regression models, the proportion of the sample included was ∼90% of the total sample; specific numbers included in each analysis is provided below the regression tables (tables 4 and 5).

**Table 1.** Summary of NHS CHECK Surveys used in this analysis by time point.

| Survey | Timepoint (T) | Launched | Closed | Key milestones in COVID-19 pandemic in UK [21] |
| --- | --- | --- | --- | --- |
| Baseline | 1 | April 2020 | January 2021 | 5 weeks after initial lockdown |
| Six-month follow-up<sup>a</sup> | 2 | October 2020 | August 2021 | UK enters second national lockdown (Nov 2020) |

*Note. The survey launch date was fixed, but the start date for each NHS Trust was staggered based on their agreement to participate.*
<sup>a</sup>*Individuals were contacted with survey invites six months after they completed the baseline survey.*

**Table 2.** Demographic characteristics and workplace experiences of participants (N= 9,769).

|  | Frequency (n) | Weighted (%) |
| --- | --- | --- |
| Exposure Variables (T1) |  |  |
| **Ethnicity** |  |  |
| White British | 7814 | 74.85 |
| White Other<sup>a</sup> | 836 | 8.01 |
| Black<sup>b</sup> | 278 | 4.79 |
| Asian<sup>c</sup> | 503 | 9.78 |
| Mixed/Other<sup>d</sup> | 285 | 2.58 |
| Missing | 53 | <1 |
| **Migration Status** |  |  |
| Born in UK | 8269 | 81.02 |
| Born in EU (Not UK) | 573 | 5.82 |
| Born elsewhere | 849 | 13.16 |
| Missing | 78 | <1 |
| Outcome Variables (T2) |  |  |
| **Support Programme Use** |  |  |
| Support Used | 8042 | 84.67 |
| Support Not Used | 1533 | 15.33 |
| Missing | 194 | 1.99 |
| **Felt Supported by Manager** |  |  |
| Little/No Support | 6944 | 69.12 |
| Supported | 2803 | 30.88 |
| Missing | 22 | <1 |
| **Felt Supported by Colleagues** |  |  |
| Little/No Support | 7924 | 81.37 |
| Supported | 1830 | 18.63 |
| Missing | 15 | <1 |
| Covariates (T1 and T2) |  |  |
| **Age (T1)** |  |  |
| ≤30 | 1452 | 17.06 |
| 31-40 | 1948 | 23.97 |
| 41-50 | 2566 | 25.52 |
| 51-60 | 2635 | 25.29 |
| ≥61 | 700 | 8.16 |
| Missing | 468 | 4.80 |
| **Sex (T1)** |  |  |
| Male | 1780 | 24.27 |
| Female | 7911 | 75.73 |
| Missing | 78 | <1 |
| **Role (T1)** |  |  |
| Doctor | 702 | 9.93 |
| Nurse | 2342 | 28.22 |
| Other clinical | 2833 | 32.28 |
| Non-clinical | 3775 | 29.56 |
| Missing | 117 | 1.20 |

<sup>a</sup> includes White Irish, White Gypsy or Irish Traveller and any other White background
<sup>b</sup> Includes Black African, Black Caribbean and any other Black background
<sup>c</sup> Includes Indian, Pakistani, Bangladeshi, Chinese, Arab or any other Asian background
<sup>d</sup> Includes any mixed background or other ethnic groups

**Table 3.** Median GHQ-12 scores for HCWs with probable CMD at T2 by ethnicity and migration status

|  | **Probable CMD<sup>ab</sup> at T2** |  |  |
| --- | --- | --- | --- |
|  | **n (%)** | **Median** | **Interquartile range** |
| **Total sample** | 4817 (51.335) | 4 | 1-8 |
| **Ethnicity** |  |  |  |
| White British | 3829 (52.49) | 4 | 1-8 |
| White Other | 444 (51.93) | 4 | 1-8 |
| Black | 117 (53.84) | 3 | 1-6 |
| Asian | 242 (40.75) | 3 | 1-8 |
| Mixed/Other | 156 (65.18) | 4 | 1-8 |
| **Migration status** |  |  |  |
| Born in UK | 4075 (52.33) | 4 | 1-8 |
| Born in EU (Not UK) | 305 (53.84) | 4 | 1-8 |
| Born elsewhere | 398 (44.24) | 3 | 1-6 |

<sup>a</sup> Common Mental Disorder
<sup>b</sup> (GHQ-12: cut-off $\geq 4$ , scale range: 0–12)
<sup>c</sup> includes White Irish, White Gypsy or Irish Traveller and any other White background
<sup>d</sup> Includes Black African, Black Caribbean and any other Black background
<sup>e</sup> Includes Indian, Pakistani, Bangladeshi, Chinese, Arab or any other Asian background
<sup>f</sup> Includes any mixed background or other ethnic groups

**Table 4.** Variations in support programme use by ethnicity and migration status.

|  | Support Used (T2) | Unadjusted |  | Adjusted<sup>a</sup> |  |
| --- | --- | --- | --- | --- | --- |
|  | n (%) | OR | 95% CI | AOR | 95% CI |
| **Ethnicity<sup>b</sup>** |  |  |  |  |  |
| White British | 6437 (84.78) | - | - | - | - |
| White Other<sup>d</sup> | 686 (84.38) | 0.87 | 0.70-1.08 | 0.85 | 0.66-1.10 |
| Black<sup>e</sup> | 227 (84.29) | 0.86 | 0.68-1.08 | 0.97 | 0.65-1.45 |
| Asian<sup>f</sup> | 412(84.70) | 0.87 | 0.47-1.63 | 0.85 | 0.45-1.60 |
| Mixed/Other<sup>g</sup> | 236 (87.15) | 1.10 | 0.65-1.87 | 1.00 | 0.56-1.78 |
| **Migration Status<sup>c</sup>** |  |  |  |  |  |
| Born in UK | 6825 (84.83) | - | - | - | - |
| Born in EU (Not UK) | 471 (86.94) | 1.10 | 0.72-1.68 | 0.97 | 0.62-1.52 |
| Born elsewhere | 681 (83.63) | 0.81 | 0.62-1.06 | 0.85 | 0.63-1.14 |

<sup>a</sup>Model adjusted for age, sex, role and GHQ-12 score at T2 and reported as AOR.
<sup>b</sup>n=9447 included in the unadjusted analysis, n=8784 included in the adjusted analysis.
<sup>c</sup>n=9421 included in the unadjusted analysis, n=8759 included in the adjusted analysis.
<sup>d</sup> Includes White Irish, White Gypsy or Irish Traveller and any other White background
<sup>e</sup> Includes Black African, Black Caribbean and any other Black background
<sup>f</sup> Includes Indian, Pakistani, Bangladeshi, Chinese, Arab or any other Asian background
<sup>g</sup> Includes any mixed background or other ethnic groups

**Table 5.** Variations in feeling supported by managers and colleagues by ethnicity and migration status.

| Exposure Variables (T1) | Manager Support at T2 |  |  | Colleague Support at T2 |  |  |
| --- | --- | --- | --- | --- | --- | --- |
|  | Felt Supported<br>n (%) | AOR<sup>a</sup> | 95% CI | Felt Supported<br>n (%) | AOR<sup>a</sup> | 95% CI |
| **Ethnicity<sup>b</sup>** |  |  |  |  |  |  |
| White British | 5597 (69.41) | - | - | 6408 (82.50) | - | - |
| White Other<sup>d</sup> | 585 (67.83) | 0.95 | 0.76-1.19 | 656 (78.35) | **0.79** | **0.64-0.99** |
| Black<sup>e</sup> | 202 (71.76) | 0.95 | 0.63-1.41 | 225 (80.26) | 0.71 | 0.50-1.03 |
| Asian<sup>f</sup> | 339 (68.92) | 0.93 | 0.74-1.16 | 384 (77.33) | **0.65** | **0.57-0.74** |
| Mixed/Other<sup>g</sup> | 192 (62.24) | 0.79 | 0.45-1.39 | 216 (76.79) | 0.82 | 0.61-1.09 |
| **Migration Status<sup>c</sup>** |  |  |  |  |  |  |
| Born in UK | 5942 (68.99) | - | - | 6787 (81.96) | - | - |
| Born in EU (Not UK) | 391 (63.53) | 0.85 | 0.60-1.20 | 452 (79.24) | 0.86 | 0.72-1.02 |
| Born in Other | 609 (71.84) | 1.09 | 0.80-1.49 | 679 (78.38) | **0.70** | **0.52-0.94** |

<sup>a</sup>Model adjusted for age, sex, role and GHQ-12 score at T2 and reported as AOR.
<sup>b</sup>n=8937 included in the adjusted analysis for manager support and n=8943 for the adjusted analysis for colleague support.
<sup>c</sup>n=8912 included in the adjusted analysis for manager support and n=8918 for the adjusted analysis for colleague support.
<sup>d</sup>Includes White Irish, White Gypsy or Irish Traveller and any other White background
<sup>e</sup>Includes Black African, Black Caribbean and any other Black background
<sup>f</sup>Includes Indian, Pakistani, Bangladeshi, Chinese, Arab or any other Asian background
<sup>g</sup>Includes any mixed background or other ethnic groups

### Socio-Demographic Characteristics

In total, 9,769 HCWs completed both the baseline and six-month follow-up questionnaire (short version). The response rate for the ‘Mixed Other’ group was similar at 89%. Response rates for the other ethnic groups was lower: ‘Black’ (81%), ‘Asian’ (82%) and ‘White Other’ (88%). Similarly, the response rate for HCWs who were born in the EU (87%) and elsewhere (84%) was lower than those born in the UK (89%).

Within ethnic groups, HCWs from the White Other ethnic group had the highest proportion of participants from migrant groups (79%) (proportion of HCWs from migrant groups by ethnicity can be found in supplementary material D).

### Common Mental Disorders and Support Use

As shown in Table 3, over half of the participants met the cut-off (score of four or more) on the GHQ-12 at T2 (51%), with a median score of 4 and an interquartile range (IQR) of 1 to 8, indicating that over half of the participants had symptoms of a probable CMD.

Table 3 shows the prevalence of a probable CMD and median GHQ-12 score across ethnic and migrant groups. The proportion of HCWs from the Mixed/Other ethnic groups (65%) was higher than the proportion of HCWs in the White British (52%) ethnic group with a probable CMD at T2. Of those HCWs born in the EU, 54% had a probable CMD at T2, slightly higher than HCWs born in the UK (52%). Conversely, of HCWs born elsewhere, 44% had a probable CMD at T2 which is lower (44%) than HCWs born in the UK (52%).

### Support Programme Use (Variations in Ethnicity and Migration Status)

In total, 8042 (85%) reported using one or more support programme (Table 2). Table 4 shows the proportion of staff who used support programmes by ethnicity and migration status. The multi-level binary logistic regression models (Table 4) showed no evidence support programme use varied by ethnicity or migration status.

### Feeling Supported (Variations by Ethnicity and Migration Status)

#### Managers

In total, 6944 (69%) HCWs reported feeling supported by a manager at T2. The multi-level binary logistic regression models (Table 5) showed no evidence that feeling supported by a manager at T2 varied by ethnicity or migration status.

#### Colleagues

In total, 7924 (81%) HCWs reported feeling supported by colleagues at T2. The multi-level binary logistic regression models (Table 5) showed HCWs from the White Other ethnic groups (AOR 0.79; CI 0.64-0.99) and HCWs from the Asian ethnic groups (AOR 0.65; CI 0.57-0.74) were less likely than HCWs from the White British ethnic group to feel supported by their colleagues at T2 after adjustment. The regression models (Table 5) showed HCWs who were born somewhere other than the UK and EU (AOR 0.70; CI 0.52-0.94) were less likely than HCWs born in the UK to feel supported by their colleagues at T2 after adjustment.

## DISCUSSION

Using a sample of NHS CHECK participants who had completed the baseline survey (launched in April 2020) and the six-month follow-up, our findings indicated that HCWs from the White Other and Asian ethnic groups were less likely than HCWs from the White British ethnic group to feel supported by their colleagues at the six-month follow-up. Furthermore, HCWs born outside the UK or EU were less likely to feel supported by their colleagues at six-month follow-up than those born in the UK. There was no variation in HCW’s use of formal support programmes or managerial support by ethnicity or migration status.

### Findings in Relation to Previous Research

Existing research highlighted high levels of bullying, harassment, and discrimination experienced by HCWs from ethnic minority and migrant groups. Despite this, little is known about their support from peers and managers during the COVID-19 pandemic or their use of formal support services. This study aimed to address this critical gap and identify disparities in staff support.

Most study participants reported having used at least one support programme at six-month follow-up (85%) a very similar finding to another study which examined rates of support use amongst UK HCWs at the beginning of the pandemic (84%) [12]. This may be at least partially explained by our finding of a substantially higher proportion (52%) of the sample having a probable CMD compared to UK working adults (24%) during the early part of the pandemic (April to November 2020) [28].

Previous general population research found that Black African, Asian and Mixed ethnic groups were less likely to self-refer to NHS Talking Therapies (formerly referred to as IAPT) than those from the White British ethnic group [29]. Additionally, there was low uptake of the Wellbeing Hubs from Black, Asian and Mixed/Other ethnic minority groups during the COVID-19 pandemic [13]. Encouragingly, in our sample, HCWs from these groups were no less likely to use support programmes than those from the White British ethnic group.

However, study participation from HCWs from Black and other ethnic minority groups was low; it is thus possible that participants with more negative experiences are less likely to participate in the surveys. In addition, the previous research focused only on psychological support, whilst our study included all support types.

Conversely, regarding informal support, our findings show that HCWs from the White Other and Asian ethnic groups were less likely than their peers from the White British ethnic group to feel supported by their colleagues.In addition, HCWs born outside of the UK were less likely to feel supported by their colleagues. This is perhaps unsurprising given the levels of abuse from other staff towards HCWs from ethnic minority groups [8]. It also mirrors a review of the experiences of international nurses who reported feeling devalued by the UK nursing establishment and having poor working relationships [30].

### Strengths and Limitations

A detailed and more nuanced exploration of ethnic variations was limited due to small participant numbers. We needed to use broad categories such as ‘White Other’ and ‘Black’ which encapsulated HCWs from various communities with distinct cultures and experiences. However, data were weighted according to each participating NHS Trust’s workforce’s demographic composition, meaning the results are more representative of those populations. Furthermore, due to the large heterogeneity of support offered by each NHS Trust, it was not feasible to explore different types of support in this analysis. This exploration would help better understand variations in access. For example, practical support such as food discounts are likely to have different motivations. There may be additional barriers and stigma associated seeking psychological support compared to practical support. Therefore, this is something we hope to explore in the future.

In addition, there was no time frame in the support use question, so it is impossible to know when staff used a support programme. The association between staff support use and mental health could include individuals who have enduring mental health difficulties and have used support over many years rather than being reflective of the impact of the COVID-19 pandemic on support needs.

As noted above, response rates for ethnic minority staff in our survey were lower than White British HCWs. HCWs from ethnic minority groups are generally underrepresented in health research [31]. Therefore, future research should use strategies to encourage participation from these groups.

Despite these limitations, this analysis was conducted on a relatively large sample size. It was derived from a wider data set that contains comprehensive information on staff of all role types (clinical and non-clinical) across urban and rural regions across England, allowing for generalisability of findings to all HCWs.

### Implications

HCWs from White Other ethnic groups were less likely than HCWs from the White British ethnic group to feel supported by their colleagues. The latest WRES report found HCWs from White Gypsy or Irish Traveller ethnic groups experienced the highest level of staff abuse [32]. Although there were very few individuals from White Gypsy or Irish Traveller ethnic groups in our study and so this may not explain our finding, it does highlight that further disaggregation of the ‘White Other’ group is valuable. The WRES has previously not distinguished between White British HCWs and those who would be in our defined ‘White Other’ ethnic group, such as individuals born in Europe. Given our findings, we encourage the WRES to continue detailed analysis of White ethnic sub-groups to better understand these disparities.

HCWs from the White Other and Asian ethnic groups and those born outside of the UK/EU were less likely to feel supported by colleagues than White British HCWs and those born in the UK, respectively. These findings together are perhaps unsurprising given that the White Other and Asian ethnic groups had the highest proportion of staff born outside of the UK (79% and 65%). Previous qualitative evidence suggests the intersectionality of ethnicity, migration status and other characteristics, such as gender, influence HCWs’ experiences of reduced agency and alienation at work [33]. Due to small numbers, we could not assess the interaction between ethnicity and migration status, highlighting the need for further analysis to explore the role of intersectionality and collegial support.

Although there were some disparities in colleague support, there was no evidence of disparities in perceived support from managers. Compassionate and supportive managers can enable individuals to use health and wellbeing workplace support services [4].

Therefore, it is a positive finding that there were no disparities in managerial support by ethnicity or migration status. Nonetheless, a significant proportion of our sample (29%) reported feeling unsupported by a manager. Training managers to confidently discuss health and wellbeing with their staff could increase the perceived level of support and it was an ambition of the NHS People Plan 2020-21 [34] that every member of the NHS should have a health and wellbeing conversation and a personalised plan. An evaluation of brief active listening skills training for managers was found to increase the confidence of managers to support distressed colleagues [35]. In addition, a case study from the Staffordshire and Stoke on Trent Care Commissioning Groups showed a decrease in sickness absence from three per cent to less than one per cent four months after the implementation of staff wellbeing conversations in team meetings [36].

## Conclusion

In conclusion, whilst it is encouraging that we did not find significant variation in the use of support programmes or managerial support by ethnicity or migration status, the critical disparities in collegial support we identified are of concern. Given the protective impact that colleague support can have against crises like the COVID-19 pandemic and ongoing systemic issues in the NHS. Future research should focus on furthering our understanding of the mechanisms driving workplace disparities for HCWs from ethnic minority and migrant backgrounds.

## Data Availability

Data are available on reasonable request submitted to the corresponding author.

## ACKNOWLEDGEMENTS

The study team would like to acknowledge the invaluable contribution of our Patient and Public Involvement group, collaborators and co-investigators. Without the continued support and expert knowledge of this group, this research would not have been possible. We are especially grateful to all the participants who took part in the study.

## Ethical Approval

This project is a secondary data analysis using data from the NHS CHECK Study and the Inequalities survey. Ethical approval for the NHS CHECK study was granted by the Health Research Authority (reference: 20/HRA/2107, IRAS: 282686).

## Data Access

Data are available on reasonable request submitted to the corresponding author.

## Author Contribution

BC: conceptualisation, methodology, formal analysis and writing of the original draft.

SAMS: writing – review and editing and funding acquisition.

DL: writing – review and editing and funding acquisition

RB: project administration and funding acquisition.

JO: writing – review and editing.

BD: writing – review and editing.

PA-M: writing – review and editing.

ZC: writing – review and editing.

NG: writing – review and editing.

CW: writing – review and editing.

R Raine: writing – review and editing

SH: conceptualisation, methodology, writing – review and editing, supervision and funding acquisition.

R Rhead: conceptualisation, methodology, writing – review and editing and supervision.

## Open Access

For the purposes of open access, the author has applied a Creative Commons Attribution (CC BY) licence to any Accepted Author Manuscript version arising from this submission.

