## Supplementary material for "Inequalities in NHS Staff Support among those from Ethnic Minority and Migrant groups during the COVID-19 Pandemic": Supplemantry Files

**Supplementary Material A.**

Table 1.

Participants in the NHS Check Study by NHS Trust

| Participating NHS Trusts | No. of participants | | ~ % of workforce participated |
| --- | --- | --- | --- |
| Avon and Wiltshire Mental Health Partnership NHS Trust | | 979 | 22.6% |
| Cornwall Partnership NHS Foundation Trust | | 1093 | 27.5% |
| Cambridgeshire and Peterborough NHS Foundation Trust | | 1495 | 35.3% |
| Cambridge University Hospitals NHS Foundation Trust | | 1017 | 9.0% |
| Devon Partnership NHS Trust | | 1386 | 42.2% |
| East Suffolk and North Essex NHS Foundation Trust | | 447 | 4.4% |
| Gloucestershire Hospitals NHS Foundation Trust | | 731 | 8.6% |
| Guys and St Thomas' NHS Foundation Trust | | 3044 | 15.4% |
| King's College Hospital NHS Foundation Trust | | 2053 | 15.8% |
| Lancashire and South Cumbria NHS Foundation Trust | | 852 | 13.4% |
| Norfolk and Norwich University Hospitals NHS Foundation | | 1376 | 13.1% |
| Nottinghamshire Healthcare NHS Foundation Trust | | 2522 | 28.5% |
| Royal Papworth Hospital NHS Foundation Trust | | 153 | 7.3% |
| Sheffield Health and Social Care Trust | | 485 | 18.6% |
| South London and Maudsley NHS Foundation Trust | | 1550 | 30.1% |
| Tees Esk and Wear Valleys NHS Foundation Trust | | 1980 | 27.1% |
| University Hospitals of Derby and Burton NHS | | 503 | 3.8% |
| University Hospitals of Leicester NHS Foundation Trust | | 1680 | 10.1% |
| Total | | **23, 346** |  |

*Note. This table is up to date as of 07.12.21 and may fluctuate depending on the addition and withdrawal of participants.*

**Supplementary Material B**

Multi-level generalised linear models showed that HCWs who used support had an estimated 0.35 unit increase in their GHQ-12 score at T2 compared to those who did not use support at T2 (OR 0.35; 95% CI 0.24 to 0.46). Secondly, HCWs who felt supported by their manager at T2 had a 1.13 unit decrease in their GHQ-12 total score at T2 compared to those who did not feel supported (OR -1.13; 95% CI -1.41 to -0.86). Finally, HCWs who felt supported by their colleagues at T2 had a 1.03 unit decrease in their GHQ-12 total score at T2 compared to those who did not feel supported at T2 (OR -1.03; 95% CI -1.21 to -0.85).

Table 1.

Associations between support use, manager support and colleague support and score on the GHQ-12.

| **Model** | **Coefficient** | **P-Value** | **95% CI** |
| --- | --- | --- | --- |
| **Support Used** | 0.35 | <0.001 | 0.24 - 0.46 |
| **Manager Support** | -1.13 | <0.001 | -1.41 - -0.86 |
| **Colleague Support** | -1.03 | <0.001 | -1.21 - -0.85 |

**Supplementary Material C**

Table 1.

The proportion of HCWs included in the analysis by the original 17 ethnic groups

| **Ethnic Group** | **Frequency (n)** | **Weighted (%)** |
| --- | --- | --- |
| White English/Welsh/Scottish/Northern Irish | 7814 | 75.00 |
| White Irish | 183 | 1.67 |
| White Gypsy or Irish Traveller | * | <1 |
| Any other White background | 649 | 6.12 |
| Mixed White and Black Caribbean | 53 | <1 |
| Mixed White and Black African | 30 | <1 |
| Mixed White and Asian | 64 | <1 |
| Any other Mixed/Multiple ethnic background | 77 | <1 |
| Indian | 240 | 4.59 |
| Pakistani | 44 | <1 |
| Bangladeshi | 27 | <1 |
| Chinese | 46 | <1 |
| Any other Asian background | 133 | 2.73 |
| Black African | 149 | 2.64 |
| Black Caribbean | 112 | 1.98 |
| Any other Black background | 17 | <1 |
| Arab | 13 | <1 |
| Any other ethnic group | 61 | 1.48 |
| **Total** | 9716 | 100 |

**Supplementary Material D**

Table 1.

Proportion of Staff Born in UK or elsewhere by Ethnicity

| **Ethnicity** | **Born in UK**  **n (%)** | **Born outside of UK^a^**  **n (%)** | ***Total***  ***n (%)*** |
| --- | --- | --- | --- |
| White British | 7561 (96.87) | 216 (3.13) | *777 (100)* |
| White Other | 178 (20.78) | 657 (79.22) | *835 (100)* |
| Black | 142 (48.70) | 136 (51.30) | *278 (100)* |
| Asian | 185 (33.65) | 316 (66.35) | *501 (100)* |
| Mixed/Other | 190 (52.56) | 95 (47.44) | *285 (100)* |
| ***Total***  ***n (%)*** | *8256 (81.24)* | *1420 (18.76)* | *9676* |

^a^These categories were combined due to small cell counts.
